# Prevalence of Malaria in Tanzania: A Systematic Review and Meta-Analysis

**DOI:** 10.64898/2026.01.15.26344205

**Authors:** Hillary R. Sebukoto, Charles B. Kafaiya, Angelina M. Lutambi, Paul M. Hayuma, Johnson J. Mshiu, Obadia Bishoge, Irene R. Mremi, Kunda J. Stephen, Jonathan M. Mshana, Mary T. Mayige

**Affiliations:** National Institute for Medical Research (NIMR), Tanga centre, Tanzania; National Institute for Medical Research (NIMR), HQ-Sub office, Dar es Salaam, Tanzania; National Institute for Medical Research (NIMR), Dodoma centre, Tanzania; National Institute for Medical Research (NIMR), Muhimbili centre, Tanzania

**Keywords:** Malaria prevalence, Systematic review, Meta-analysis, *Plasmodium falciparum*, Tanzania

## Abstract

**Background:** Understanding malaria epidemiology in Tanzania is a step toward achieving malaria control and elimination targets by 2030. This meta-analysis derived robust estimates of malaria prevalence and *Plasmodium* species from studies in Tanzania published from 2014 – 2024.

**Methods:** Following the PRISMA guidelines, we searched PubMed, Google Scholar, and Science Direct using the PICOS framework for study selection. We used the Simonian-Laird random-effects model to obtain the pooled malaria prevalence. Study quality was assessed using Covidence, with heterogeneity quantified by the I^2^ index.

**Results:** We included 31 high-quality studies. Pooled malaria prevalence was 17% (95% CI: 11–22, I² = 98.1%, p < 0.001) by microscopy, 19% (95% CI: 12–25, I² = 99.8%, p < 0.0001) by RDT, and 8% (95% CI: 2–14, I² = 94.8%, p < 0.001) by PCR. Children had the highest rates: 22% (RDT), 19% (microscopy), and 20% (PCR); adults had 11%, 14%, and 36%, respectively. Common mixed infections involved *P. falciparum* with *P. ovale* (22%) or *P. malariae* (18%). P. vivax co-infections were rare (5%). Lake zone regions had the highest disease prevalence (36.5%).

**Conclusion:** Malaria disproportionately affects children in Tanzania, underscoring their vulnerability to infection. The predominance of *P. falciparum* mixed infections underscores the complex epidemiology in the country. The high burden in Lake zones emphasizes the need for targeted interventions. Enhanced diagnostics, focused control strategies, and surveillance are essential in reducing the disease burden and advancing control efforts in the country.

## Introduction

Malaria remains a major global public health challenge, with cases rising to 249 million and 608,000 deaths in 2022, a nearly 6% increase since 2019 [1–3]. The African region bears 94% of global cases and 95% of all deaths [3–5]. Factors such as climate change and disruptions during the COVID-19 pandemic have exacerbated the burden threatening progress in malaria control [6–10]. Among other sub-Saharan African countries, Tanzania contributes significantly to the global number of cases and deaths [11,12].

Malaria transmission involves symptomatic infections, which prompt treatment and limit spread, versus asymptomatic cases that persist undetected as hidden reservoirs sustaining community transmission. These carriers challenge elimination efforts, requiring sensitive diagnostics to break infection cycles and support public health interventions [13,14].

In Tanzania, where malaria remains a leading cause of morbidity and mortality across all age groups [15,16], understanding symptomatic and asymptomatic burdens is crucial. National surveys and hospital records indicate that the disease accounts for a substantial proportion of outpatient visits and hospital admissions [17,18]. Despite the reductions in malaria cases and deaths over the past two decades, lake and coastal regions experience persistently high transmission rates [19,20]. This is compounded by the fact that approximately 93% of Mainland Tanzania’s population resides in transmission areas [12], with patterns varying across regions [21,22]. Studies have reported wide prevalence estimates, influenced by differences in populations, locations, and diagnostic [23–27]. This heterogeneity underscores the need for data synthesis to precisely assess the national burden

Several studies across Tanzania’s regions have estimated malaria prevalence [24,27–29], but yield conflicting results due to variation in scope, methodology, sample sizes, and diagnostics. This meta-analysis pools these data for precise national estimates, identifying high-burden and vulnerable groups needing intensified interventions, and reveals demographic/regional disparities to guide evidence-based control and resource allocation. Understanding this burden evaluates intervention effectiveness, informs policy, and supports strategic planning; thus, this systematic review and meta-analysis consolidates evidence to provide robust prevalence estimates, laying a foundation for context-specific research and public health efforts toward reducing malaria morbidity and mortality

## Materials and Methods

### Settings

This review included studies conducted in the United Republic of Tanzania, an East African country. Tanzania shares borders with eight countries and the Indian Ocean and has an estimated population of about 62 million people as per the 2022 National Census [30]. The country’s diverse geography and climatic zones contribute to substantial spatial variations in malaria transmission.

### Review procedures

The systematic review was conducted in accordance with the Preferred Reporting Items for Systematic reviews and Meta-Analyses (PRISMA) guidelines, from defining the research question to manuscript preparation, providing a structured framework for conducting and reporting systematic reviews, encompassing all sections of the manuscript, including the title, abstract, introduction, methods, results and discussion [31–33].

### Protocol registration

The protocol for this systematic review and meta-analysis was registered through PROSPERO with an ID number PROSPERO 2025 CRD420251006620.

### Literature search and search strategy

The Preferred Reporting Items for Systematic Reviews and Meta-Analyses (PRISMA) methodology was used for the literature search, study selection, data extraction, and reporting of findings for this review [34]. Comprehensive searches were conducted in PubMed, Google Scholar, and ScienceDirect to identify articles on malaria prevalence across all Tanzania regions. We used a methodical keyword search approach, matching search phrases to Medical Subject Headings (MeSH). Boolean operators (AND, OR) were used to combine or separate keywords to improve search precision. The search combined MeSH terms (“Prevalence”[MeSH], “Malaria”[MeSH], “Plasmodium”[MeSH], “Plasmodium falciparum”[MeSH]) AND location terms covering Tanzania[MeSH] plus all regions in Tanzania. All studies on the prevalence of malaria in Tanzania that were published between 2014 and 2024 were included.

### Selection criteria and search outcomes

The PICOS (Population, Intervention, Comparison, Outcomes, and Study Design) model was used to define the study selection criteria. The population included all humans, regardless of age or sex. As the focus was on prevalence, no specific intervention or comparison was required. The primary outcome was the reported prevalence of malaria in Tanzania, and eligible study designs included original cross-sectional studies, cohort studies, case-control studies, experimental studies, or modelling studies. The review also included studies reporting the prevalence of malaria in Tanzania over the past 10 years (2014–2024). The following exclusion criteria were applied: animal studies or laboratory-based studies without clinical relevance; reviews, letters, notes, editorials, conference reports, and qualitative studies.

The search process identified a total of 611 articles related to the review topic. All Studies were imported into Covidence, a web-based systematic review management software, for screening, full article review and data extraction. A total of 31 studies that met the set criteria were included in this review (Figure 1).

**Figure 1.**
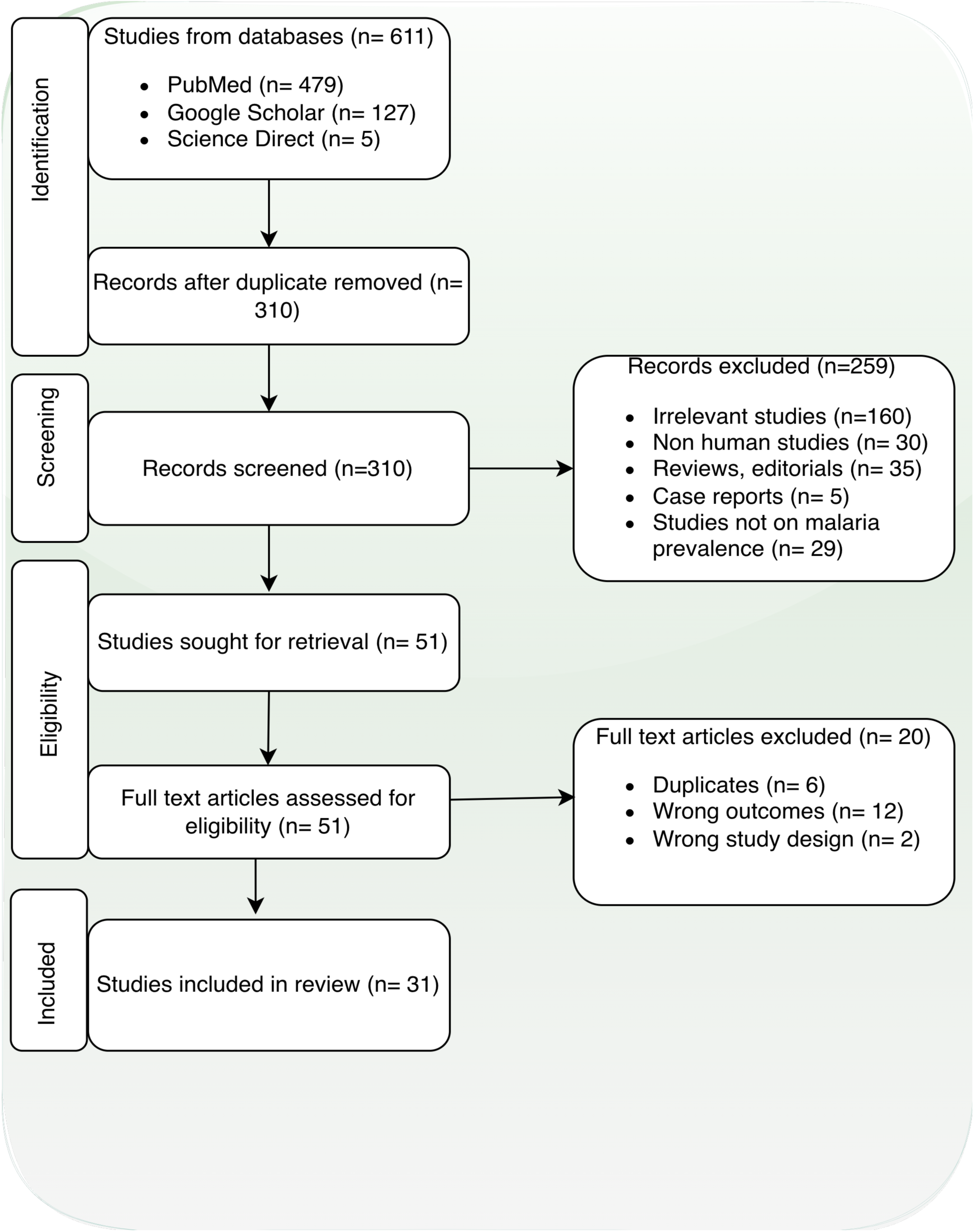
PRISMA Scheme of Retrieved Studies.

### Information sources

A comprehensive search was conducted across multiple databases and sources to identify studies reporting on the prevalence of malaria in Tanzania. The bibliographic databases search included PubMed (https://pubmed.ncbi.nlm.nih.gov/) and Google Scholar (https://scholar.google.com/), Science Direct (https://www.sciencedirect.com/), with all searches completed in March 2025. These databases were selected for their extensive coverage of public and global health literature. All sources were searched with restrictions on publication date (2014-2024), and search strategies were tailored to each database using appropriate keywords and Boolean operators.

### Quality assessment of the included studies

Quality assessment was conducted using Covidence, an online review-management platform that facilitated a structured and transparent evaluation of study bias. We assessed each study using nine criteria from the Joanna Briggs Institute (JBI) for prevalence studies, scoring each item (yes = 1; no or unclear = 0). Four independent reviewers recorded judgments and supporting evidence within Covidence, and any disagreements were resolved through consensus. The analysis showed that the included studies had a low risk of bias, with an average total score of 96%, which exceeds the 50% threshold defined for acceptable quality (Table 1).

**Table 1.**
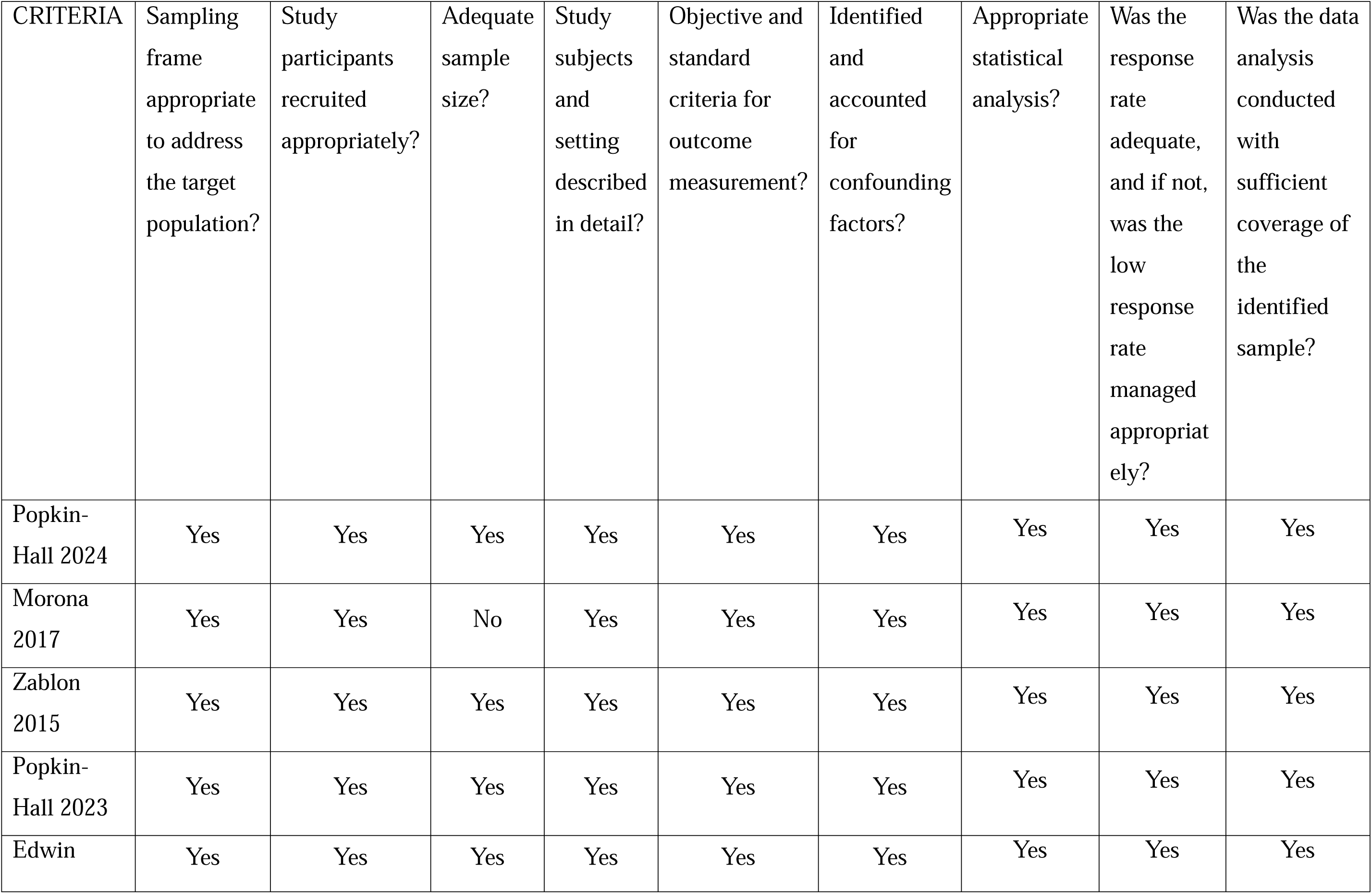
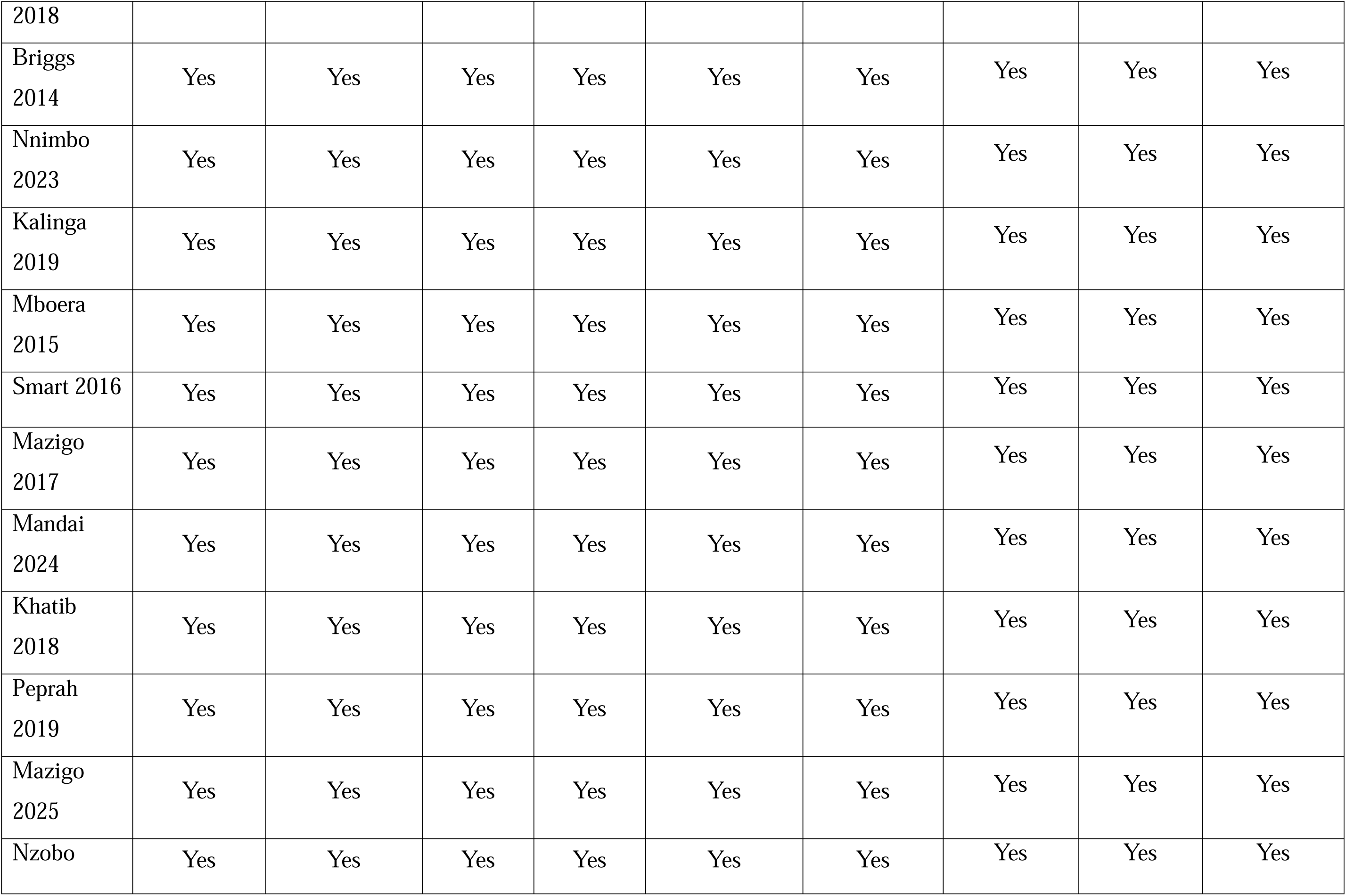
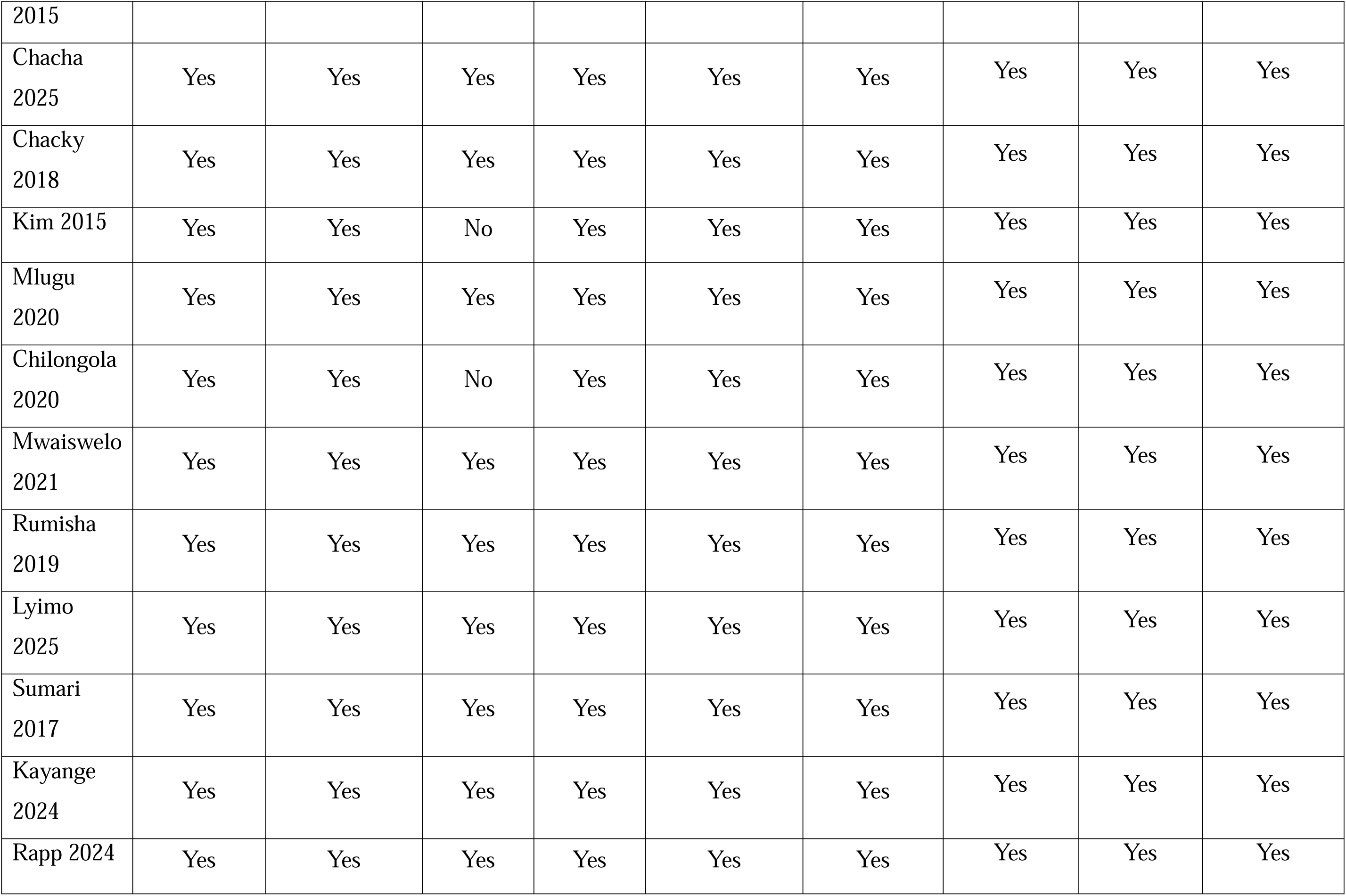
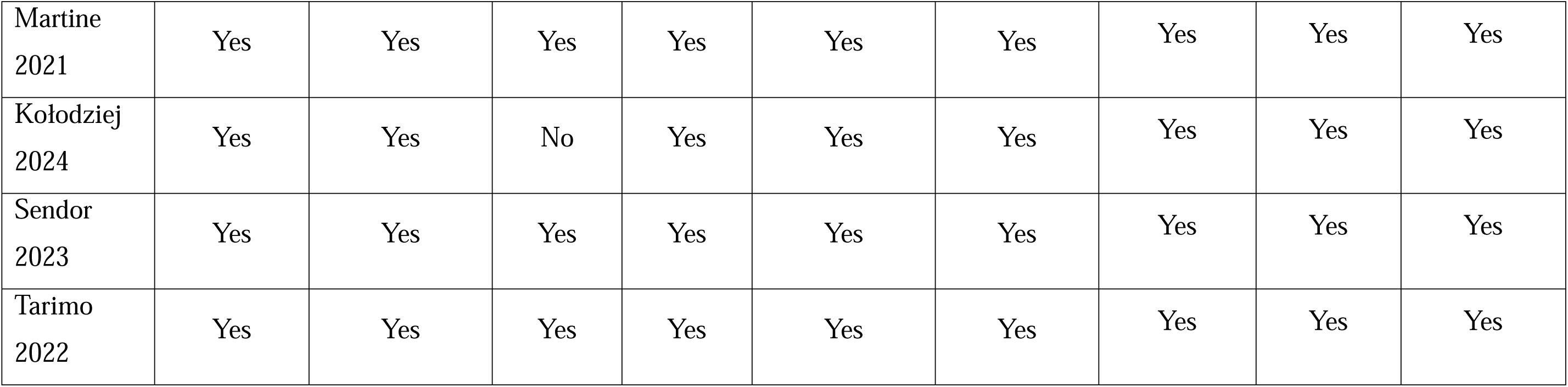
Assessment of Study Quality and Potential Bias in Included Studies.

### Data extraction, data analysis and synthesis

The following data were extracted from each included article: study ID, study title, study DOI, authors, study location, study objective, design, start and end dates, population description, inclusions and exclusion criteria, participants’ recruitment methods, sample size, baseline population characteristics and malaria prevalence proportions by species and overall.

After extracting the data via Covidence data extraction sheet, we utilized Stata MP. 17 version for statistical analysis. Because of the considerable heterogeneity in the study, we adopted random-effects models. Otherwise, it generates study encumbrances, mostly used to distinguish between different research variations. The I^2^ statistic, which has a value ranging from 0 to 100%, predicts that the major difference among studies involved in a systematic review and meta-analysis is attributable to heterogeneity instead of chance. Low, medium and high heterogeneity between trials is indicated by I^2^ values of 25%, 50% and 75% plus. When the p-value was less than 0.05, the presence of heterogeneity was determined.

### Study selection

The search strategy initially identified 611 titles for screening. After eliminating duplicates, 310 titles and abstracts were screened. After a full-text review of 51 studies, 31 met our inclusion criteria. There was substantial agreement between four reviewers at the titles and abstract screening stage (κ = 0.781; 95% CI, 0.749-0.812) and at the full-text screening stage (κ =0.702; 95% CI, 0.581-0.822) (Figure1).

### Patient and public involvement (PPI)

Patients and the public were not involved in the design, conduct, or reporting of this study, as it is a systematic review and meta-analysis of published cross-sectional studies using only secondary aggregate data, making direct PPI not applicable

## Results

### Study characteristics

A total of 31 cross-sectional studies conducted across various regions of Tanzania were included in this review. The studies covered diverse geographical areas, including both mainland and coastal regions. About 45.2% of the studies targeted participants of all age groups, while others focused specifically on school-aged children, under-fives, adults, and pregnant women. The studies were published between 2014 and early 2025, reflecting a growing research interest in malaria across Tanzania. Overall, the included studies provide a comprehensive representation of malaria prevalence across different demographic groups and ecological settings within the country (Table 2).

**Table 2.** Characteristics of the included studies.

| Study | Region | Study population |
| --- | --- | --- |
| Briggs et al., 2014(35) | Mwanza and Mtwara | All ages |
| Chacha et al., 2025(36) | Kagera, Kigoma, Njombe, Ruvuma and Tanga | All ages |
| Chacky et al., 2018(26) | Tanzania Mainland | School children |
| Chilongola et al., 2020(37) | Tanga | All ages |
| Edwin et al., 2018(38) | Tanzania Mainland | Under-fives |
| Kalinga et al., 2019(39) | Kigoma, Tanga, Ruvuma and Mara | Adults |
| Kayange et al., 2024(40) | Mwanza | Children |
| Khatib et al., 2018(41) | Pwani | All ages |
| Kim et al., 2015(42) | Mwanza | School children |
| Kołodziej et al., 2024(43) | Arusha | All ages |
| Lyimo et al., 2025(44) | Tanga | Children |
| Mandai et al., 2024(45) | Kagera | All ages |
| Martine et al., 2021(46) | Tanga | Children |
| Mazigo et al., 2017(20) | Morogoro | All ages |
| Mazigo et al., 2025(47) | Geita, Kigoma and Arusha | All ages |
| Mboera et al., 2015(48) | Morogoro | School children |
| Mlugu et al., 2020(49) | Pwani | Pregnant women |
| Morona et al., 2017(50) | Mwanza | Adults |
| Mwaiswelo et al., 2021(51) | Mtwara | All ages |
| Nnimbo et al., 2023(52) | Tabora | Adults |
| Nzobo et al., 2015(27) | Morogoro | School children |
| Peprah et al., 2019(53) | Simiyu, Geita, Mara & Mwanza | Children |
| Popkin-Hall et al.,<br>2023(54) | Dar es Salaam, Dodoma, Kagera, Kilimanjaro,<br>Manyara, Mara and Mtwara | All ages |
| Popkin-Hall et al.,<br>2024(55) | Ruvuma, Tanga and Kigoma | All ages |
| Rapp et al., 2024(56) | Pwani | All ages |
| Rumisha et al., 2019(19) | Morogoro | All ages |
| Sendor et al., 2023(57) | Arusha, Iringa, Kagera, Mara, Mtwara,<br>Rukwa, Tabora and Tanga | School children |
| Smart et al., 2016(58) | Mwanza | Children |
| Sumari et al., 2017(28) | Pwani | School children |
| Tarimo et al., 2022(59) | Pwani | All ages |
| Zablon et al., 2015(60) | Dodoma | Pregnant women |

### Epidemiological Profile of Malaria Prevalence

Out of 31 studies included in this review, 15 reported specifically on malaria prevalence. Using a random-effects inverse-variance model, the pooled prevalence of malaria was estimated at 16% (95% CI: 12–21%). Subgroup analysis by diagnostic method revealed pooled prevalence estimates of 19% (95% CI: 12–25%) for rapid diagnostic tests (RDTs), 8.0% (95% CI: 2–14%) for polymerase chain reaction (PCR), and 17% (95% CI: 11–22%) for microscopy, with all estimates statistically significant (p < 0.05). There were substantial heterogeneity across studies (overall I² = 99.6%, p < 0.001), with significant differences between subgroups (p = 0.037).

The Prevalence varied significantly by age group and diagnostic method. Children exhibited the highest rates, with 22% detected by RDT, 20% by PCR, and 19% by microscopy, whereas adults had lower rates of 11%, 36%, and 14%, respectively. Overall, RDTs consistently produced higher positivity rates (25%) compared to microscopy (16%) and PCR (18%) (Figure 2).

**Figure 2.**
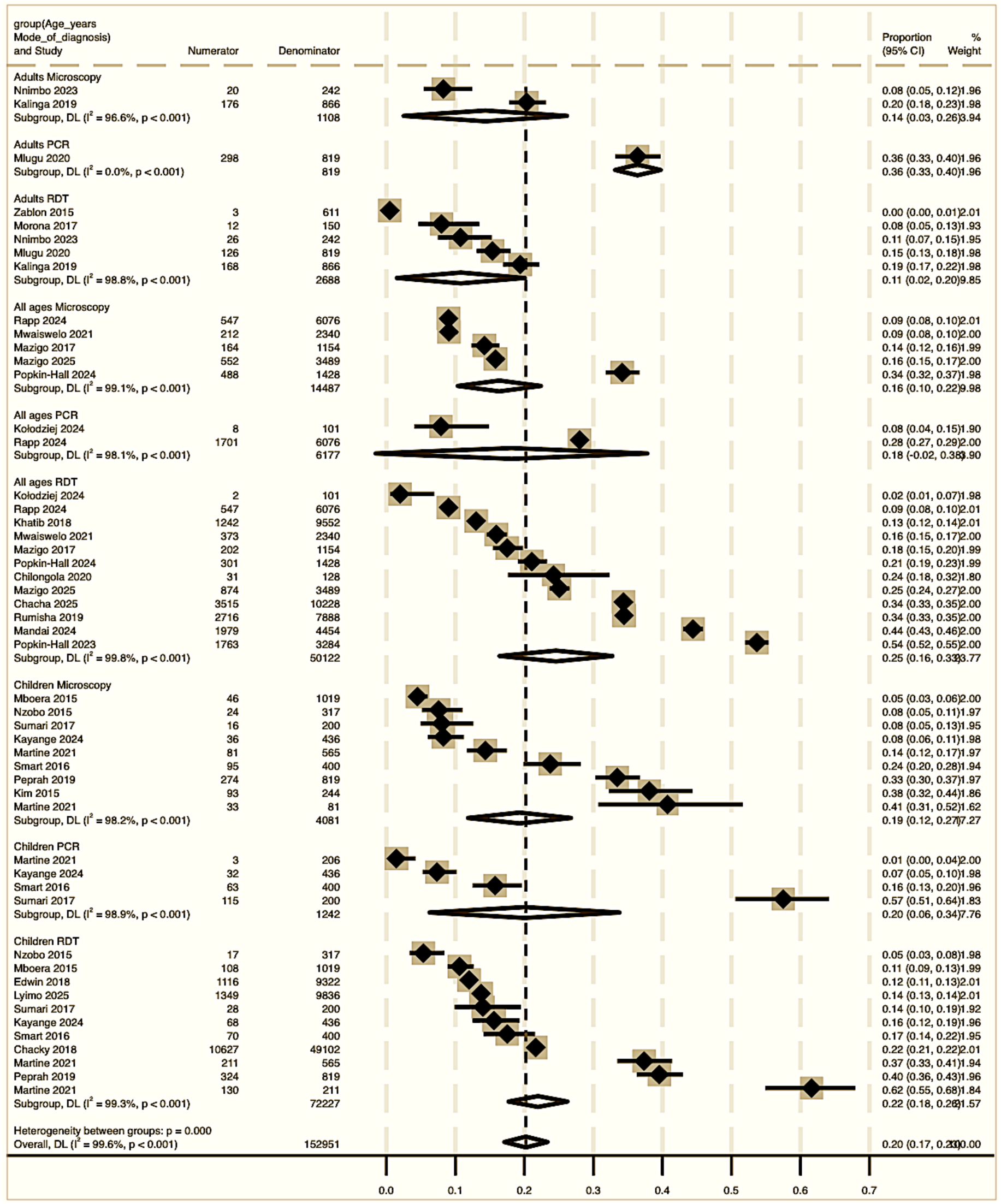
Pooled Prevalence of Malaria in Tanzania (2014-2024), Stratified by Age Group and Diagnostic Mode.

For asymptomatic malaria, pooled prevalence also varied by diagnostic method using a random-effects inverse-variance model. RDTs showed prevalence ranging from 5% to 62%, with a pooled estimate of 22% (95% CI: 15–28%); microscopy-based estimates ranged from 8% to 41%, yielding 19% (95% CI: 12.0–25.0%); and PCR-based prevalence ranged from 28.0% to 57.0%, with 40% (95% CI: 28.0–52%). All subgroup estimates were statistically significant (p < 0.001), but substantial heterogeneity persisted (RDTs I² = 99.7%, microscopy I² = 98.8%, PCR I² = 97.7%, overall, I² = 99.6%, p < 0.001). Between-subgroup analysis indicated significant differences by diagnostic method (p = 0.008). P. falciparum dominated asymptomatic infections, with non-falciparum species like P. malariae and P. ovale detected at lower frequencies. PCR-based studies revealed greater species diversity than microscopy or RDTs. Mixed-species infections involved P. falciparum with P. malariae (18%) or P. ovale (22%), and rarely P. vivax (5%) (Figure 3).

**Figure 3.**
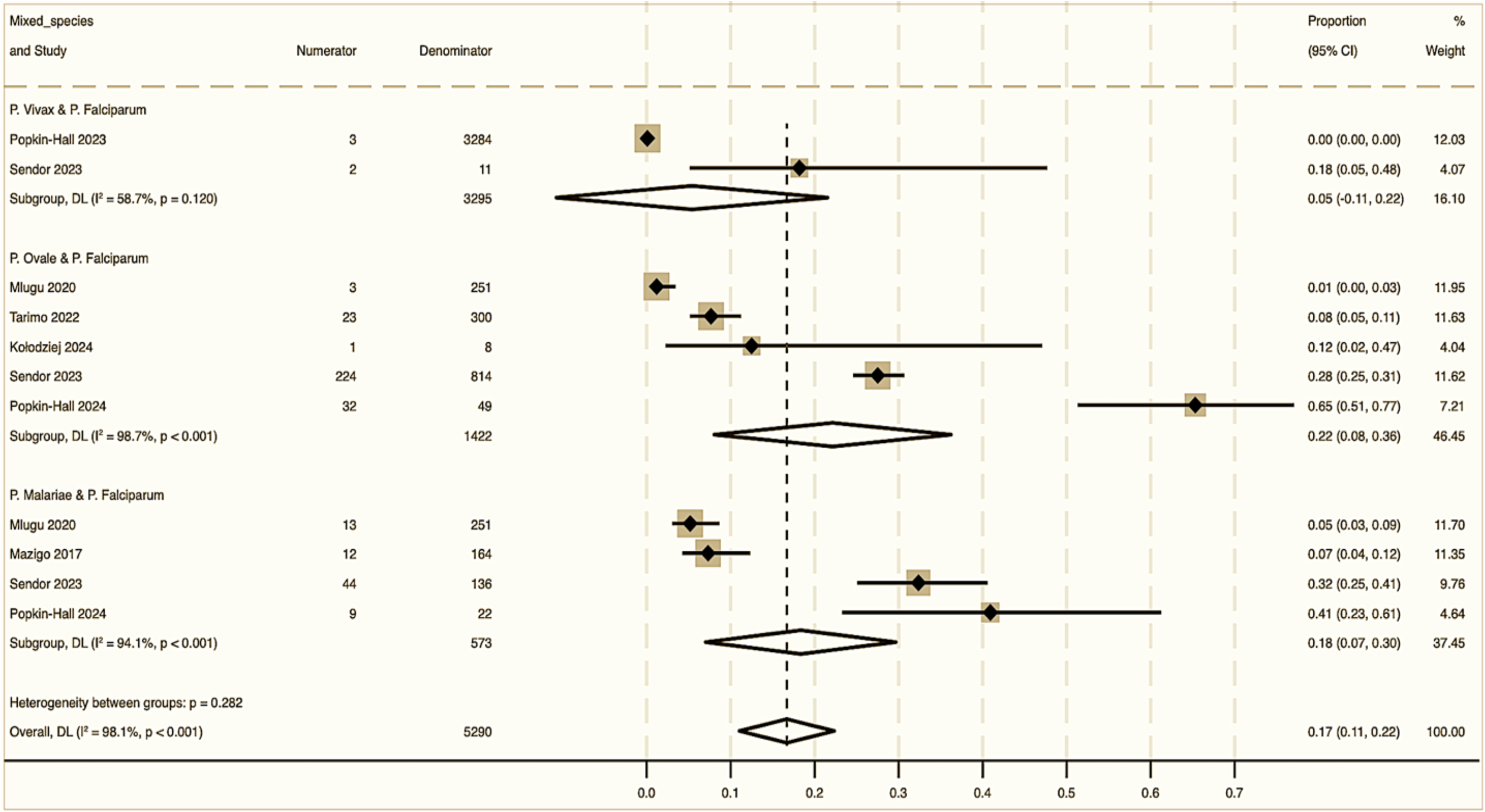
Prevalence of Malaria by Mixed Species (2014-2024)

Notable geographical variations emerged, with the highest prevalence in the Lake Zone (Simiyu, Geita, Mara, and Mwanza; 18–36.5%), followed by Coastal regions (Tanga 18.2%, Mtwara 12.5%) and Western Zone (Tabora 9.5%). Lower rates occurred in Northern and Central zones (Arusha 5.0%, Dodoma 0.5%).

### Publication bias

The observed asymmetry from the funnel plot may suggest small-study effects or publication bias, though it could also represent genuine heterogeneity due to variations in diagnostic methods, settings, or populations across studies. The significant heterogeneity (Q = 6603.43) necessitates cautious interpretation of the asymmetry.

## Discussion

The significant malaria burden in Tanzania is highlighted by this systematic review and meta-analysis, which is in line with earlier research and reports from several African nations, including Nigeria, Uganda, Kenya and Ethiopia [61–64]. The regional similarity may not only be due to shared ecological conditions, but also to environmental conditions, which include high vector density, favourable breeding sites, and comparable socio-economic and health system factors influencing malaria transmission [65–67]. These findings have important implications for malaria diagnostics, surveillance, and control efforts. The choice of diagnostic method influenced the prevalence estimates, with rapid diagnostic tests (RDTs) consistently providing the highest rates, while PCR-based diagnostics reported the lowest rates (19% and 8%, respectively). The observed variation reflects underlying differences in test sensitivity, antigen persistence with RDTs, and the detection thresholds specific to each assay [68]. Furthermore, the observed higher prevalence with microscopy compared to PCR might be due to operator skill and variability in parasite densities, as reported in different literature [69,70]. These results underscore malaria as a significant public health concern and suggest that achieving the malaria elimination goal in Tanzania by 2030 will necessitate a shift to more targeted, evidence-based strategies that are informed by comprehensive surveillance and the strategic selection of diagnostic tools.

Age-specific analysis indicates that malaria prevalence is significantly higher in children than in adults, aligning with WHO findings of greater susceptibility in younger populations [5]. This increased risk is due to incomplete immunity and frequent mosquito exposure [71,72]. Variations were noted in the prevalence of *Plasmodium falciparum*, with RDTs showing potential underestimation of malaria cases in children. Similar studies in Nigeria, Uganda, and Ethiopia also found that RDT sensitivity is lower compared to microscopy in detecting malaria in children, particularly in areas with lower transmission rates [73–75].

The prevalence of asymptomatic malaria detected by PCR (40%) significantly exceeds that detected by microscopy (19%) and RDT (22%). This aligns with research indicating a higher sensitivity of PCR for low-density infections compared to conventional methods, which often miss submicroscopic carriers [76]. While microscopy is the gold standard for diagnosing malaria due to its quantification capabilities, it struggles with low parasitemia sensitivity [77]. This limitation echoes findings from Uganda [78] and highlights the risks of relying solely on such surveillance methods. RDTs, although rapid and specific, also have higher detection thresholds than PCR, potentially leading to underestimation of asymptomatic cases [79]. The substantial 40% positivity rate underscores asymptomatic malaria as a hidden reservoir for transmission, raising concerns about the sufficiency of current surveillance practices, especially in endemic regions like Tanzania, impacting future malaria control strategies.

A notably higher prevalence of mixed-species malaria infections was observed in the current meta-analysis, with a pooled prevalence of 17%, which is significantly higher compared to previous literature [80]. A meta-analysis conducted in Southern Asia reported that the risk of severe malaria among patients with mixed infections was found to be comparable to those with P. falciparum mono-infection, suggesting that there are possibilities of mixed infections to offer protective immunological effects, such as reduced risk of severe anaemia with P. falciparum/P. vivax co-infection due to cross-species immunity [81]. However, studies report that mixed infections are often under-recognized by standard microscopy, which can lead to inadequate treatment and an increased risk of drug resistance [82]. This is a salient challenge, especially in Tanzania, where there is evidence of artemisinin partial resistance in the northwestern part of the country [45]. This, therefore, complicates the management of mixed infections, as inadequate or misdiagnosed treatment could further spread resistant strains, suggesting a need for enhanced molecular surveillance and tailored malaria control programs in the country [83].

Pronounced regional heterogeneity in malaria prevalence was observed, representing one of the study’s most important findings. The Lake Zone, particularly Simiyu, exhibited the highest prevalence (36.5%), confirming it as a persistent hotspot. In comparison, Dodoma reported a very low prevalence of 0.5%, highlighting substantial geographical disparities despite national control efforts. It is possible to hypothesize that ecological factors, vector density, and breeding conditions contribute to these regional differences [22,84]. These findings highlight the persistent heterogeneity of malaria transmission across ecological zones, with lake and coastal areas remaining the country’s primary malaria hotspots.

These data must be interpreted with caution because of the substantial heterogeneity observed across all subgroups in the meta-analysis. It may be the case that some low-prevalence estimates detected by PCR reflect limitations in detecting low-density infections rather than true reductions in transmission. Despite these limitations, this review provides comprehensive and valuable insights into malaria prevalence patterns, age-specific burdens, and regional disparities, offering critical guidance for spatially targeting future control interventions.

## Conclusion

This systematic review reveals marked heterogeneity in malaria prevalence across Tanzania’s mainland regions, underscoring the imperative for geographically tailored interventions. Established best practices warrant sustained implementation, while evidence gaps demand urgent attention. Robust surveillance systems paired with highly sensitive diagnostics remain essential to attaining elimination targets by 2030

## Acknowledgement

The authors would like to express their sincere gratitude to the researchers who made their work publicly available online; their contributions were instrumental in making this review possible.

## Author’s contribution

Conceptualization, HRS, CBK, PMH, AML, MTM, JMM, KJS, IRM, OB and JJM; Protocol registration, Literature search, Full article review, Writing the manuscript, HRS, CBK, MTM, AML and PMH; Data analysis and Interpretation, AML and CBK; Screening, Data extraction and Review, HRS, PMH and CBK; Review and editing of the manuscript, MTM, JMM and AML

## Funding statement

This research received no specific grant from any funding agency in the public, commercial or not-for-profit sectors.

## Informed consent statement

Not applicable.

## Data availability

The datasets used in this review are available upon request to the corresponding author.

## Competing interests

No conflict of interest declared.

## Notes

### Competing Interest Statement

The authors have declared no competing interest.

### Funding Statement

This study did not receive any funding

### Summary of Updates

Have updated the name of the corresponding author, as well as rearranging a list of authors

